# Association of Cancer History with Structural Brain Aging Markers of Alzheimer’s Disease and Dementia Risk

**DOI:** 10.1101/2023.02.19.23286154

**Authors:** Jingxuan Wang, Kendra D. Sims, Sarah F. Ackley, Ruijia Chen, Lindsay C. Kobayashi, Eleanor Hayes-Larson, Elizabeth Rose Mayeda, Peter Buto, Scott C. Zimmerman, Rebecca E. Graff, M. Maria Glymour

## Abstract

**Background and Objectives:** Cancer survivors are less likely than comparably-aged individuals without a cancer history to develop Alzheimer’s disease and related dementias (ADRD). We investigated the association between cancer history and structural magnetic resonance imaging (MRI) markers for ADRD risk, using linear mixed-effects models to assess differences at the mean values of MRI markers and quantile regression to examine whether the association varies across the distribution of MRI markers of brain aging.

**Methods:** Among UK Biobank participants with ≥1 brain MRI, we considered total gray matter volume, total brain volume, hippocampal volume, white matter hyperintensity volume, and mean cortical thickness in the Alzheimer’s disease (AD) signature region. Cancer history was ascertained from national registry and self-report. We first specified linear mixed models with random intercepts to assess mean differences in MRI markers according to cancer history. Next, to examine whether effects of cancer history on these markers varies across the ADRD risk distribution, we specified quantile regression models to assess differences in quantile cut-points of the distribution of MRI markers according to cancer history. Models adjusted for demographics, APOE-ε4 status, and health behaviors.

**Results:** The sample included 42,242 MRIs on 37,588 participants with no cancer history (mean age 64.1 years), and 6,073 MRIs on 5,514 participants with a cancer diagnosis prior to MRI (mean age 66.7 years). Cancer history was associated with smaller mean hippocampal volume (b=-19 mm^3^, 95% confidence interval [CI]=-36, -1) and lower mean cortical thickness in the AD signature region (b=-0.004 mm, 95% CI=-0.007, -0.000). Quantile regressions indicated cancer history had larger effects on high quantiles of white matter hyperintensities (10^th^ percentile b=-49 mm^3^, 95% CI=-112, 19; 90^th^ percentile b=552 mm^3^, 95% CI= 250, 1002) and low quantiles of cortical thickness (10^th^ percentile b=-0.006 mm, 95% CI=-0.011, -0.000; 90^th^ percentile b=0.003 mm^3^, 95% CI=-0.003, 0.007), indicating individuals most vulnerable to ADRD were more affected by cancer history.

**Discussion:** We found no evidence that cancer history was associated with less ADRD-related neurodegeneration. To the contrary, adults with cancer history had worse MRI indicators of dementia risk. Adverse associations were largest in the highest-risk quantiles of neuroimaging markers.

## Introduction

Numerous epidemiologic studies have identified an inverse association between both prevalent and incident diagnosis of cancer of any type and subsequent risk of incident Alzheimer’s disease and related dementias (ADRD).^1–4^ Some studies have also found that individuals with a history of cancer had lower neuropathologic burden of Alzheimer’s disease (AD) compared to age-matched decedents without a history of cancer.^5–7^ Possible explanations for the observed inverse association include selective survival of healthier cancer survivors who are lower risk of ADRD and shared biological or genetic mechanisms that elevate cancer risk but reduce ADRD risk.^8–12^ The inverse association between cancer and ADRD has sparked substantial interest because biological explanations, if validated, could offer important insights into the underlying biology of ADRD. While existing research has focused on incident ADRD or cognitive function as the outcome of interest, there is a lack of evidence on the relationship between cancer history and biomarkers of ADRD risk. Understanding of this relationship could help to disentangle potential biases (e.g., selective survival bias, diagnostic bias) from causal mechanisms in the cancer-ADRD relationship.^8^

Measuring ADRD onset is difficult due to the slow etiologic development of the disease, phenotypic heterogeneity, and frequent missed or delayed diagnoses.^13–15^ Brain changes detectable from neuroimaging likely precede diagnosed ADRD by decades.^16^ Evaluating whether cancer history is associated with neuroimaging markers of ADRD risk avoids the potential for diagnostic bias of ADRD which occurs when clinicians overlook ADRD symptoms due to cancer diagnoses and treatments or when patients with a cancer history have more frequent contact with clinicians, increasing chance of ADRD diagnosis. While there is research on treatment-induced structural and functional brain changes among breast cancer survivors,^17–19^ only two prior studies have evaluated the association between cancer diagnosis and MRI markers of structural brain aging; the sample sizes in both studies were too small (n=2,043 and 1,609) to provide precise estimates for individual cancer types.^20,21^

In this manuscript, we used the UK Biobank neuroimaging sample to test the hypothesis that people with a history of cancer have brain MRI characteristics associated with lower risk of incident ADRD compared to otherwise similar individuals without a history of cancer. We also evaluated heterogeneous effect of cancer across quantiles of each MRI marker because individuals most vulnerable to ADRD may also be affected the most by cancer history.

## Methods

### Study population

The UK Biobank is a prospective volunteer cohort of 502,490 adults aged 40-69 years who attended one of 22 assessment centers across the United Kingdom from 2006-2010. At the baseline visit, participants completed physical, physiological, and medical assessments. In 2014, the UK Biobank invited participants for brain MRIs at three clinics using identical protocols. At the time of writing, MRI data were available for 43,102 participants, among whom 5,514 had data from a repeated imaging visit.

### Brain imaging data

Neuroimaging variables were selected *a priori* based on previous studies showing their associations with cognitive decline or ADRD pathologies. They included total grey matter volume,^22,23^ total brain volume,^24^ hippocampal volume,^25,26^ white matter hyperintensity volume,^27,28^ and mean cortical thickness in the AD signature region, comprising six regions of interest: entorhinal, inferior temporal, middle temporal, inferior parietal, fusiform, and precuneus.^29^ All MRIs were carried out using similar scanners (Siemens Skyra 3T scanner with a standard 32-channel head coil). Full details on image acquisition, processing, and quality control are available from the UK Biobank Brain Imaging Documentation (https://biobank.ctsu.ox.ac.uk/crystal/crystal/docs/brain_mri.pdf) and protocol publications.^30^ In brief, the T1-weighted anatomic images were acquired using three-dimensional magnetization prepared for rapid gradient-echo (3D MPRAGE) at a resolution of 1 × 1 × 1 mm. Volumetric measures were calculated with the FreeSurfer ASEG. Total white matter hyperintensity volumes were derived based on T1 and T2 fluid-attenuated inversion recovery (FLAIR) using the Brain Intensity Abnormality Classification Algorithm (BIANCA).^31^ Regional estimates of cortical thickness and surface area were processed using FreeSurfer v.5.3 based on the Desikan–Killiany atlas parcellation.^32^ Hippocampal volume was estimated by summing left and right hippocampal volumes. All volumetric measures were corrected for skull size using a residual method^33^ that calculated adjusted brain volumes from residuals of a linear regression between raw volumes and intracranial volume. The mean cortical thickness in the AD signature region was calculated by the surface area-weighted average of cortical thicknesses across six regions of interest.

### Ascertainment of cancer cases

Cancer diagnoses were identified via linkage to hospital admission, cancer registries, and self-reported medical conditions using International Classification of Diseases ICD-10 and ICD-9 codes (Supplementary Table 1). We included all cancer types except non-melanoma skin cancer (NMSC) in our primary analysis. For all participants, an indicator for cancer diagnosis denoted at least one diagnosis any time prior to the first imaging visit. A time-updated indicator of cancer diagnosis was available for 5,514 participants who had a repeat imaging visit. In secondary analyses, we repeated the variable construction for breast cancer ^2,34^ and prostate cancer,^2,35^ which are the two most common cancers in the UK Biobank and have been linked to ADRD risk in previous studies. In addition, evidence of gray matter volume loss and white matter microstructural disruption has been reported among breast cancer survivors treated with chemotherapy.^17,18^ To account for the potential impact of chemotherapy-induced brain changes, we also evaluated NMSC, for which chemotherapy is not a common treatment.^36^

### Ascertainment of ADRD cases

ADRD cases were identified from linkage to hospital admission, primary care, and death records using a comprehensive list of ICD-10 and ICD-9 codes described elsewhere.^9^ In brief, we included AD, vascular dementia, frontotemporal dementia, Lewy body dementia, alcohol-related dementia, and Creutzfeldt-Jakob disease. We defined ADRD onset as the first date of ADRD diagnosis.

### Assessment of covariates

We controlled for covariates that plausibly influence both cancer and ADRD risk. We conceptualized two sets of covariates based on their temporality relative to cancer diagnosis (Supplementary Figure 1). Our ‘base model’ included covariates that could not be affected by cancer diagnosis: age (linear and quadratic terms^37–39^), sex (female, male), race (White, Black, Asian, Other), and binary APOE-ε4 carrier status. In our fully adjusted models, we additionally controlled for covariates that could both affect and be affected by cancer diagnosis: educational attainment (professional/university degree, secondary, vocational, other qualifications), Townsend deprivation index, body mass index (BMI), ever smoking, ever alcohol use, high physical activity (≥75 min/week of vigorous activity or ≥150 min/week of moderate activity^40^), and assessment center (Cheadle, Reading, Newcastle, Bristol). Most covariates were assessed at study enrollment from 2006-2010. Information on BMI, smoking, alcohol use, and physical activity was collected during the imaging visits. Townsend deprivation index is a composite score measuring area-level socioeconomic status based on employment, home ownership, car ownership, and household overcrowdedness.^41^ During each visit, trained staff measured height and weight, and BMI was derived dividing weight (kg) by the square of height (m^2^). Participants reported details on ever smoking, ever alcohol use, and physical activity via touchscreen questionnaires.

### Statistical analyses

We summarized characteristics of the study sample stratified by cancer status. To confirm the relevance of the selected MRI measures for ADRD, we used Cox proportional hazards regression models to estimate adjusted hazard ratios (HR) and 95% confidence intervals (CI) for associations between each MRI marker and incident ADRD. For these analyses, we treated the imaging visit as baseline and only included participants with no ADRD diagnosis at that time. Participants were followed up to date of ADRD diagnosis, death, or censoring on September 30, 2021 (the latest date of ADRD diagnosis in the imaging cohort), whichever came first. We adjusted for the same covariate sets as in the primary analyses. We scaled brain volume and cortical thickness measures by dividing them by the sample standard deviation and estimated separate models for each individual measure.

To evaluate whether mean values of the neuroimaging outcomes differed by cancer status, we used linear mixed-effects models with individual-level random intercepts to account for repeated within-person MRI measures. To examine whether the effects of a prior cancer diagnosis on neuroimaging outcomes varied across the distribution of ADRD risk, we specified quantile regression models at the 10th, 25th, 50th, 75th, and 90th percentiles of each neuroimaging outcome. We used cluster bootstrapping to account for repeated within-person measures.

Quantile regression allows estimation of the relationship between an exposure and an outcome across cut-points of the outcome distribution, such as quartiles or deciles.^42^ The quantile regression coefficients quantify how much the location each specified quantile of the neuroimaging outcome distribution differs between those with and without a history of cancer. If the coefficients are similar in each of the five quantile regression models (10th, 25th, 50th, 75th, and 90th percentiles), we would expect the associations of cancer history with each neuroimaging outcome are not differential across the entire distribution of the outcome and vice versa. We used the Wald test to check for heterogeneous associations of cancer with neuroimaging outcomes across quantiles. In secondary analyses, we analyzed the associations of breast cancer, prostate cancer, and NMSC with mean neuroimaging outcomes. For each individual cancer type, we excluded participants with any other cancer diagnosis.

We conducted three sensitivity analyses. First, to minimize potential short-term effects of chemotherapy, we excluded participants with a cancer diagnosis within 5 years before the MRI scan. Second, to model potential time-varying associations of cancer with neuroimaging outcomes, we split the exposure variable into four categories based on the time between the most recent cancer diagnosis and the MRI scan: 0 – 1 year, >1 – 5 years, >5 – 10 years, and >10 years. Third, we applied inverse probability weighting (IPW) ^43^ to address potential selection bias^44^ due to selection into the imaging cohort from the parent UK Biobank study. We used a logistic regression model to calculate the inverse probability of selection into the imaging cohort and then assigned stabilized weights to each participant and performed inverse probability weighted analyses to address informative selection. Model specifications and weights assessment are presented in Supplemental Data 1.

All statistical analyses were performed using R version 4.0.5.

## Results

### Participant characteristics

The analytical sample included 43,102 participants (Table 1). The mean age at the first imaging visit was 64.5 years (SD = 7.7 years). Before the imaging visit, 5,514 (12.8%) participants had a recorded cancer diagnosis. Participants with a history of cancer were older, more likely to be female and White, and averaged less physical activity.

**Table 1.** Demographic, clinical, and imaging characteristics of participants in the analytical sample at the first imaging visit, stratified by cancer history.

| Characteristic | No cancer diagnosis<br>(n = 37,588) | Cancer diagnosis<br>(n = 5,514) |
| --- | --- | --- |
| Age (year), median<br>(interquartile) | 64.5 (58.1-70.1) | 67.7 (61.4-72.4) |
| Sex |  |  |
| Female | 19,337 (51.4) | 3,371 (61.1) |
| Male | 18,251 (48.6) | 2,143 (38.9) |
| Race |  |  |
| White | 36,302 (96.6) | 5,397 (97.9) |
| Black | 321 (0.9) | 30 (0.5) |
| Asian | 602 (1.6) | 44 (0.8) |
| Other | 353 (0.9) | 43 (0.8) |
| APOE-ε4 carrier |  |  |
| Yes | 10,187 (72.2) | 1,462 (72.8) |
| No | 26,416 (27.8) | 3,905 (27.2) |
| Education |  |  |
| Higher | 19,496 (51.9) | 2,769 (50.2) |
| Secondary | 13,622 (36.2) | 2,029 (36.8) |
| Vocational | 2,035 (5.6) | 285 (5.2) |
| Other | 2,435 (6.5) | 431 (7.8) |
| Townsend deprivation index<sup>b</sup> |  |  |
| 1 (least deprived) | 8,959 (23.9) | 1,330 (24.2) |
| 2-4 | 23,390 (62.3) | 3,414 (62.0) |
| 5 (most deprived) | 5,205 (13.9) | 762 (13.8) |
| BMI (kg/m<sup>2</sup>), mean (SD) | 26.5 (4.4) | 26.4 (4.4) |
| Ever smoked |  |  |
| Yes | 13,697 (36.8) | 2,232 (41.0) |
| No | 23,549 (63.2) | 3,207 (59.0) |
| Ever used alcohol |  |  |
| Yes | 36,123 (96.8) | 5,292 (96.8) |
| No | 1,213 (3.2) | 173 (3.2) |
| High physical activity |  |  |
| Yes | 31,743 (85.4) | 4,570 (84.2) |
| No | 5,406 (14.6) | 857 (15.8) |
| GM (mm<sup>3</sup>), mean (SD) | 615,546 (56,068) | 605,119 (53,262) |
| TBV (mm<sup>3</sup>), mean (SD) | 1,161,750 (111,880) | 1,143,852 (106,584) |
| HV (mm<sup>3</sup>), mean (SD) | 8,079 (808) | 7,919 (783) |
| WMH (mm<sup>3</sup>), mean (SD) | 4,968 (6,498) | 6,033 (7,761) |
| CTAD (mm), mean (SD) | 2.82 (0.12) | 2.81 (0.12) |

Values are mean (SD), count (%), or median (interquartile).
Abbreviations: BMI, body mass index; SD, standard deviation; GM, gray matter; TBV, total brain volume; HV, hippocampal volume; WMH, white matter hyperintensity; AD, Alzheimer's disease; CTAD, cortical thickness in the AD signature region.
<sup>a</sup> Percentages may not sum to 100 because of rounding.
<sup>b</sup> Index quintiles, combining social class, employment, car availability, and housing.

### Associations of brain MRI variables with incident dementia

All MRI variables were significantly associated with ADRD incidence (Table 2). In the fully adjusted models, each standard deviation increase in gray matter volume (SD = 31,251 mm^3^), total brain volume (SD = 47,163 mm^3^), hippocampal volume (SD = 678 mm^3^), and mean cortical thickness in the AD signature region (SD = 0.12 mm), was associated with 64% (HR= 0.36, 95% CI= 0.29-0.44), 50% (HR= 0.50, 95% CI= 0.42-0.60), 56% (HR= 0.44, 95% CI= 0.36-0.54), and 48% (HR= 0.52, 95% CI= 0.42-0.65) decrease in the hazard of ADRD, respectively. For each SD increase in white matter hyperintensity volume (SD = 6,616 mm^3^), there was a 27% increase in the hazard of ADRD (HR= 1.27, 95% CI= 1.11-1.45).

**Table 2.** Association of neuroimaging outcomes with incident ADRD diagnosis: HRs from Cox proportional hazards regression models.

| Exposure<sup>a</sup> | N<sup>b</sup> | Demographics-Adjusted Base Model<sup>c</sup> |  | Fully Adjusted Model<sup>d</sup> |  |
| --- | --- | --- | --- | --- | --- |
|  |  | HR (95% CI) | p-value | HR (95% CI) | p-value |
| GM | 41,178 | 0.34 (0.28-0.41) | <0.001 | 0.36 (0.29-0.44) | <0.001 |
| TBV | 41,178 | 0.49 (0.41-0.58) | <0.001 | 0.50 (0.42-0.60) | <0.001 |
| HV | 41,939 | 0.41 (0.35-0.49) | <0.001 | 0.44 (0.36-0.54) | <0.001 |
| WMH | 39,870 | 1.26 (1.11-1.42) | <0.001 | 1.27 (1.11-1.45) | <0.001 |
| CTAD | 41,939 | 0.51 (0.42-0.63) | <0.001 | 0.52 (0.42-0.65) | <0.001 |

Abbreviations: ADRD, Alzheimer's disease and related dementias; HR, hazard ratio; CI, confidence interval; GM, gray matter; TBV, total brain volume; HV, hippocampal volume; WMH, white matter hyperintensity; CTAD, cortical thickness in the AD signature region.
<sup>a</sup> Each exposure was scaled by dividing values by the sample standard deviation. All volumetric measures are in mm<sup>3</sup> and cortical thickness is in mm.
<sup>b</sup> Number of participants included in the analyses per outcome.
<sup>c</sup> Adjusted for age, sex, race, and APOE-ε4 carrier status.
<sup>d</sup> Further adjusted for education and Townsend deprivation index, BMI, ever smoking, ever using alcohol, physical activity, and assessment center.

### Associations of cancer history with neuroimaging outcomes

Compared with participants without a history of cancer, participants with a history of cancer showed smaller hippocampal volume (*β* = -19 mm^3^, 95% CI = -36 to -1) and lower cortical thickness in the AD signature region (*β* = -0.004 mm, 95% CI = -0.007 to -0.000) in fully adjusted models (Table 3). There were no differences in gray matter volume, total brain volume, or white matter hyperintensity volumes between participants with and without a cancer history.

**Table 3.** Covariate-adjusted differences in mean values of neuroimaging outcomes comparing participants with versus without cancer history from linear mixed effects regression models.

| Outcome<sup>a</sup> | N<sup>b</sup> | Demographics-Adjusted Base<br>Model<sup>c</sup> |  |  | Fully Adjusted<br>Model<sup>d</sup> |  |  |
| --- | --- | --- | --- | --- | --- | --- | --- |
|  |  | Estimate | 95% CI | p-value | Estimate | 95% CI | p-value |
| GM | 41,378 | -452 | -1146 to 241 | 0.201 | -350 | -1056 to 355 | 0.330 |
| TBV | 41,378 | -293 | -1401 to 815 | 0.604 | -376 | -1510 to 758 | 0.515 |
| HV | 42,036 | -23 | -40 to -6 | 0.008 | -19 | -36 to -1 | 0.037 |
| WMH | 40,119 | 216 | 39 to 392 | 0.017 | 178 | -1 to 357 | 0.051 |
| CTAD | 42,036 | -0.004 | -0.007 to -0.001 | 0.007 | -0.004 | -0.007 to -0.000 | 0.026 |

Abbreviations: CI, confidence interval; GM, gray matter; TBV, total brain volume; HV, hippocampal volume; WMH, white matter hyperintensity; AD, Alzheimer's disease; CTAD, cortical thickness in the AD signature region.
<sup>a</sup> All volumetric measures are in mm<sup>3</sup> and cortical thickness is in mm.
<sup>b</sup> Number of participants included in each analysis. Mixed effects models with random intercepts for individuals were used to account for a small number of repeated MRIs.
<sup>c</sup> Adjusted for age, sex, race, and APOE-ε4 carrier status.
<sup>d</sup> Further adjusted for education and Townsend deprivation index, BMI, ever smoking, ever using alcohol, physical activity, and assessment center.

The distributions of each neuroimaging outcome, stratified by cancer history, are shown in Figure 1. The quantile regression results estimating associations of cancer history with the 10^th^, 25^th^, 50^th^, 75^th^, and 90^th^ percentiles of each neuroimaging outcome are shown in Figure 2 and Supplemental Table 2. No significant differences in any quantiles of total gray matter volume and total brain volume were observed. The association between cancer history and hippocampal volume did not show clear patterns across quantiles, but cancer history was associated with a significantly smaller 75^th^ percentile of the distribution of hippocampal volume (*β*_75_ = -26 mm^3^, 95% CI = -54 to -1). This point estimate was similar to the estimated association with the 10^th^ percentile (*β*_10_=-22 mm^3^, 95% CI = -52 to 13). The magnitude of the associations between cancer history and white matter hyperintensity volume were largest for the higher values of white matter hyperintensity; for example, cancer history was associated with a 250 mm^3^ larger 75^th^ percentile (95% CI = 71 to 446) and a 552 mm^3^ larger 90^th^ percentile (95% CI = 250 to 1002). Comparing participants with versus without cancer history, the adjusted differences in 10th, 25th, 50th, 75th, and 90th percentiles of the distributions of white matter hyperintensity volumes were significantly different (p-value < 0.001). For cortical thickness in the AD signature region, associations were mainly observed in the lowest and median percentiles (*β*_10_ = -0.006 mm, 95% CI = -0.011 to -0.000; *β*_50_ = -0.005 mm, 95% CI = -0.009 to -0.001), and the associations across percentiles were significantly different (p-value = 0.003).

**Figure 1.**
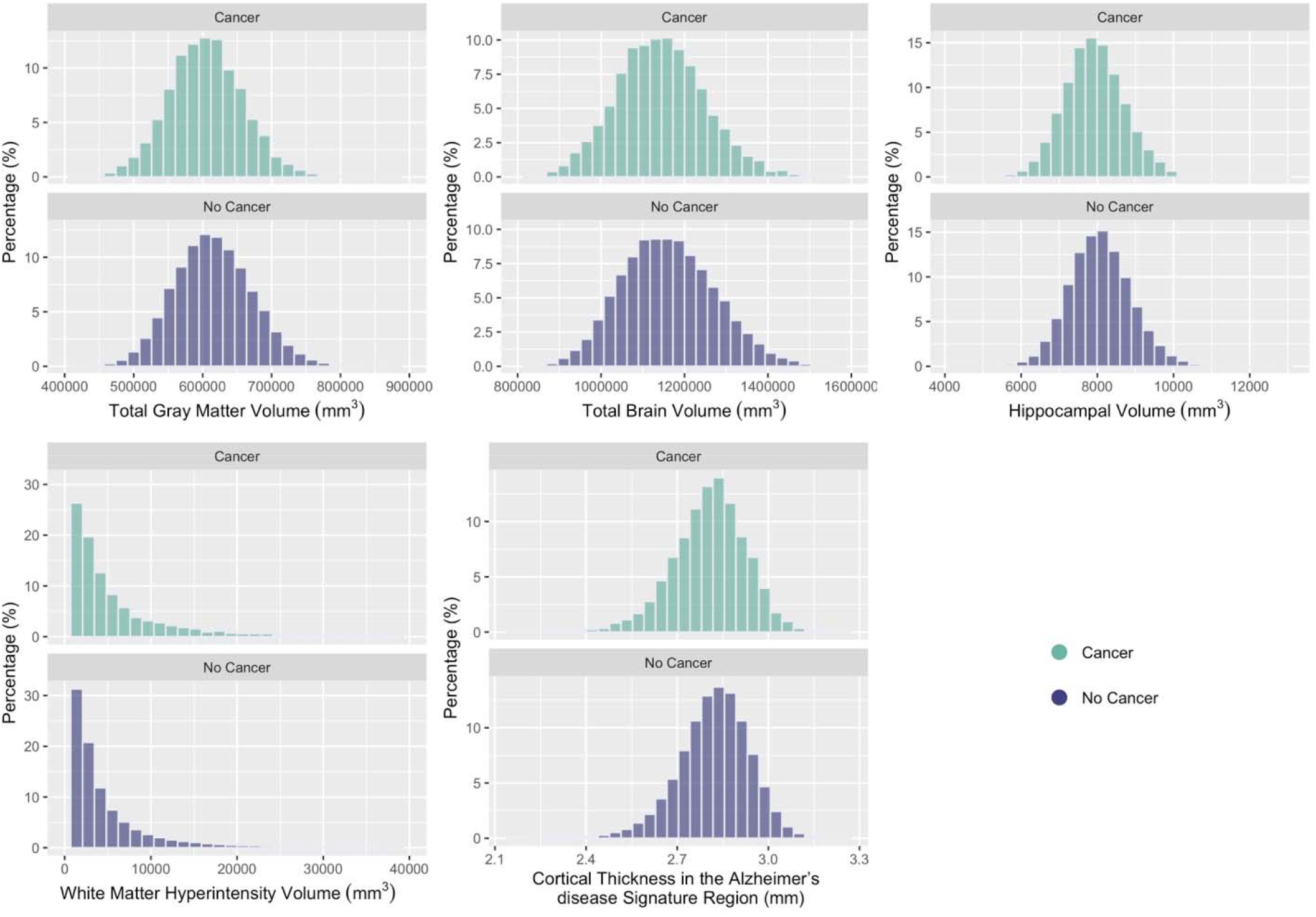
The distributions of each neuroimaging outcome, stratified by cancer history.

**Figure 2.**
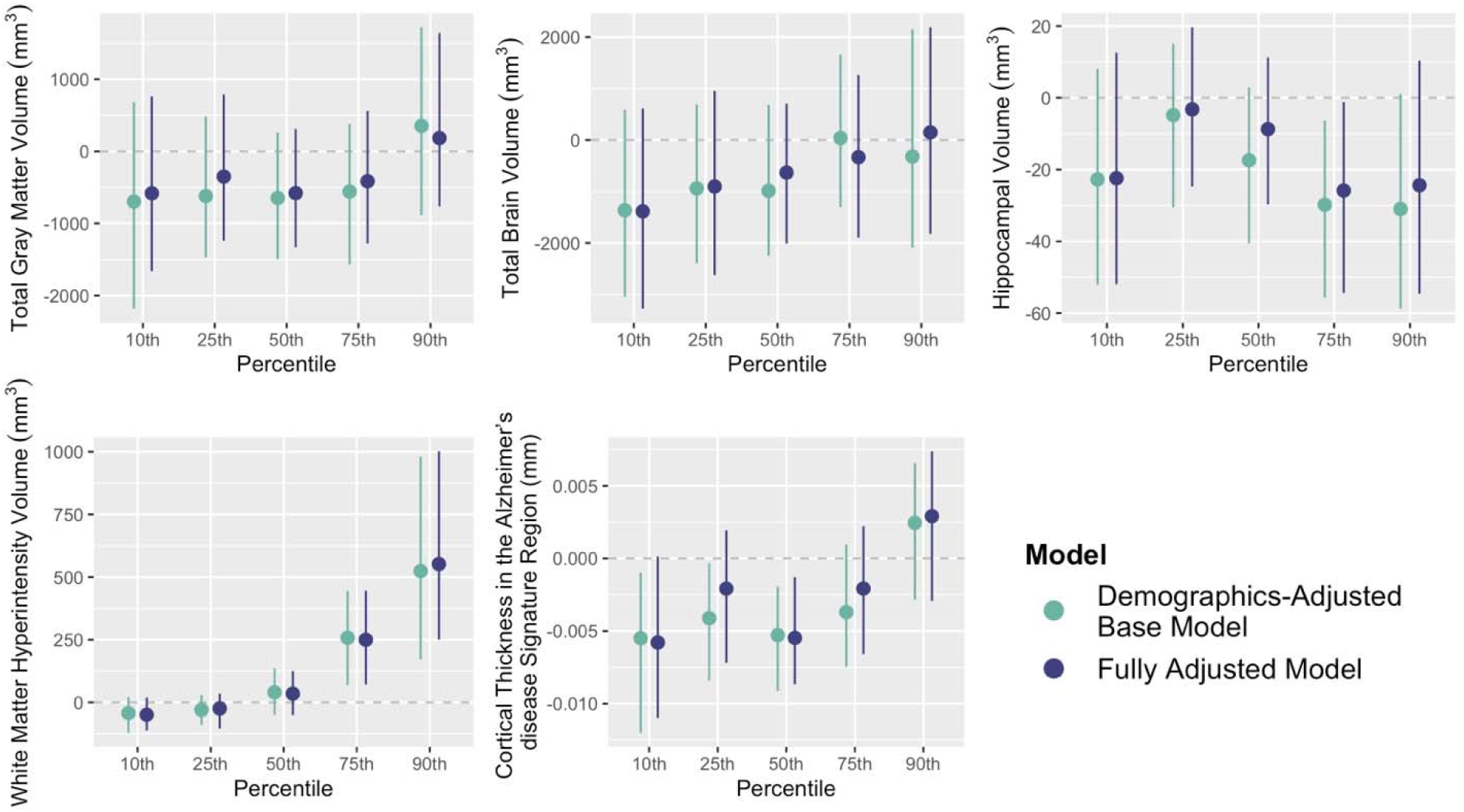
Covariate-adjusted differences in the 10^th^, 25^th^, 50^th^, 75^th^, and 90^th^ percentiles of the distributions of each neuroimaging outcome, comparing participants with versus without cancer history in quantile regression models. Note: Demographics-adjusted base model was adjusted for age, sex, race, and APOE-ε4 carrier status. Fully adjusted model was further adjusted for education and Townsend deprivation index, BMI, ever smoking, ever using alcohol, physical activity, and assessment center.

When we investigated individual common cancer types, we found no significant mean differences in any neuroimaging outcome between participants with a NMSC history and participants without any cancer history (Supplementary Table 3). Participants with a history of breast cancer compared to participants without a cancer history showed a significantly lower total brain volume (*β* = -3450 mm^3^, 95% CI = -5908 to -993), higher white matter hyperintensity volume (*β* = 512 mm^3^, 95% CI = 132 to 891), and lower cortical thickness in the AD signature region (*β* = -0.008 mm, 95% CI = -0.014 to -0.001). Prostate cancer was significantly associated with a higher total brain volume (*β* = 2832 mm^3^, 95% CI = 114 to 5551) but no other MRI markers.

### Sensitivity analyses

After we excluded participants with a cancer history within 5 years before the MRI scan, no significant differences were observed across neuroimaging outcomes (Supplementary Table 4). We did not observe clear patterns when we split the exposure based on the time between the most recent cancer diagnosis and the MRI scan, though most significant differences were detected among participants with a cancer diagnosis within 1-5 years prior to the MRI scan (Supplementary Table 5: *β* = -1516 mm^3^, 95% CI = -2797 to -236 for gray matter volume; *β* = 391 mm^3^, 95% CI = 51 to 731 for white matter hyperintensity volumes; and *β* = -0.012 mm, 95% CI = -0.018 to -0.006 for cortical thickness in the AD signature region). In addition, a cancer diagnosis within 5-10 years before the MRI scan was associated with a lower hippocampal volume (*β* = -35 mm^3^, 95% CI = -68 to -1). The inverse probability weighted analyses to estimate results if the neuroimaging sample had the same characteristics as the full UK Biobank study population did not substantially alter results (Supplementary Table 6).

## Discussion

In a large sample of UK adults, we found no evidence to support the hypothesis that individuals with a history of cancer had brain MRI markers indicative of lower ADRD risk compared to individuals without a history of cancer. Rather, adults with a history of cancer had some brain MRI markers associated with higher ADRD risk. They had slightly lower average hippocampal volume and lower cortical thickness in the AD signature region than those without a cancer history, though the sensitivity analysis suggested these findings are driven by those with a cancer diagnosis within 5 years of the MRI. Cancer history was associated with higher variability across quantiles of white matter hyperintensity volume, i.e., there were larger adverse associations with high quantiles of white matter hyperintensity volume and null associations with low quantiles of white matter hyperintensity volume. Similar patterns were seen for cortical thickness, such that the adverse association with cancer history was largest at low (high-risk) quantiles of cortical thickness. Survivors of breast cancer, an invasive cancer type, had significantly lower total brain volume and higher white matter hyperintensity volume compared to participants without a cancer history. Prostate cancer was associated with a slightly larger total brain volume. This association was not evident in NMSC.

Our results were consistent with the two previous studies describing the link between cancers at any site and structural brain aging.^20,21^ In the Framingham Heart Study (n=2,043), Gupta et al. found that compared with those without a cancer history, cancer survivors did not have a significant difference in total cerebral brain volume, temporal brain volume, temporal horn volume, or white matter hyperintensity volumes.^20^ Nudelman et al. conducted a voxel-based morphometric analysis of cerebral gray matter density (GMD) in the Alzheimer’s Disease Neuroimaging Initiative cohort (n=1,609) and did not find any region with increased GMD among cancer survivors.^21^ Instead, they found that a cancer history was associated with lower GMD in the right superior frontal gyrus. Though this area has not been linked to AD pathogenesis or diagnosis,^45^ it has been associated with cancer treatments.^46,47^ Our results expand on these findings in a sample over 10 times larger than the combined previous samples, with a wider range of neuroimaging measures, and evaluated whether effects differed across the distribution of neuroimaging outcomes. Our results indicate that, if anything, cancer history is associated with higher risk of ADRD. The quantile regression results bolster this interpretation by showing differential effects across the distribution of the outcomes. Differentially harmful effects of cancer history at higher-risk quantiles of white matter hyperintensity volume and cortical thickness suggest that individuals most at-risk for dementia may also be harmed the most by cancer history.

Our results also indicated that cancer treatment may play a role in ADRD-related neurodegeneration. In the sensitivity analysis with a 5-year washout period before the MRI scan, all adverse associations between cancer history and neuroimaging markers were attenuated and close to null. The attenuated differences among people who had survived 5+ years after diagnosis suggests that recent cancer treatment may have driven the loss in hippocampal volume and thinning in cortical thickness in the AD signature region. This is also supported by the association between breast cancer and adverse neuroimaging outcomes due to known chemotherapy-induced brain changes among breast cancer patients. Although our study did not evaluate this directly, these associations may reflect short-term effects of chemotherapy.^17,18^ The sensitivity analysis may also suggest potential selective survival that people with a cancer history further in the past who survived 5+ years may potentially have healthier brains relative to the full population of people diagnosed with cancer.

Although the inverse relationship between cancer and ADRD has been studied and reported in multiple epidemiological studies, ^1–4,8–12^ our results did not support an inverse link between cancer diagnosis and ADRD-related neurodegeneration, except a small positive association between prostate cancer and total brain volume. Potential biases in observational studies may have yielded previously observed associations. Most recent studies evaluating the cancer-ADRD link have been based on clinical diagnosis of ADRD, which may face methodological challenges, including missed, delayed, or misclassified diagnoses. For example, several simulation-based studies have suggested the importance of accounting for competing risk of death and diagnostic bias.^10,12^ Future work should address the biological mechanisms linking underlying cancer and subsequent ADRD and account for various study biases.

The lack of information on cancer treatment is a significant limitation of this study, especially given the potential effect of chemotherapy and hormone therapy on brain structures.^17,18,48^ We additionally lacked some important biomarkers, such as amyloid burden. Further, the UK Biobank is a highly selected volunteer sample with unusually high socioeconomic status and healthy individuals compared to the UK population.^49^ This selection process may bias observed associations.^50^ A major strength of our study is its measurement of cancer and structural MRI in a large cohort. The UK Biobank has a large sample size for participants with structural MRI, measured based on a uniformly high-quality image acquisition protocol. The availability of both cancer registry and self-report data on cancer history increased the validity of exposure measurement.

## Conclusion

In this large sample, we find no evidence that cancer history is associated with lower ADRD risk as measured by brain MRI markers. We find instead a suggestion of adverse effects of cancer history on some neuroimaging markers, including harmful associations for individuals already at high dementia risk. Future studies should evaluate longitudinal changes in cognition and neuroimaging markers before and after cancer treatments.

## Data Availability

All data produced are available online at https://www.ukbiobank.ac.uk/

