## Supplemental Material for "Association of Cancer History with Structural Brain Aging Markers of Alzheimer’s Disease and Dementia Risk"

**Supplemental Material Table of Contents**

### Supplemental Data 1. Model specifications for inverse probability weighting

We estimated probabilities of being in the imaging cohort from the parent UK Biobank study with a logistic regression. Covariates included cancer history before the imaging visit, age (linear and quadratic terms), sex, race, APOE-ε4 carrier status, educational attainment, Townsend deprivation index, body mass index (BMI), ever smoking, ever alcohol use, physical activity, assessment center, diabetes history, and stroke history. Then, we assigned stabilized weights to each participant and the final weights are:

$$SW_{i}=\frac{Pr(S=1)}{Pr(S=1|A,L)}$$

where

- S is the indicator for being in the imaging cohort
- A is the cancer history before the imaging visit
- L are all other covariates
- $Pr(S=1|A,L)$ is estimated with logistic regression
- $Pr(S=1)$ is the marginal probability estimated with the sample size of the imaging cohort divided by that of the UK Biobank cohort

Supplemental Figure 1. Conceptual model guiding selection of covariates to evaluate the link between cancer history and brain MRI differences related to ADRD risk. Base model covariates are established prior to cancer diagnoses. Full model covariates include time-varying factors that may influence cancer risk but may also be affected by a cancer diagnosis. The figure shows both pre-diagnosis and potentially post-diagnosis versions of these covariates, but in the data, we do not have temporal resolution to distinguish whether these covariates were established prior to a cancer diagnosis.

Abbreviations: AD, Alzheimer’s disease; ADRD, Alzheimer's disease and related dementias; BMI, body mass index; MRI, magnetic resonance imaging; SES, socioeconomic status.

### Supplementary Table 1. ICD codes for site-specific cancer

| Cancer | ICD-9 | ICD-10 |
| --- | --- | --- |
| All-cause excluding NMSC | 140-209 except 173 | C00-C96 except C44 |
| NMSC | 173 | C44 |
| Breast cancer | 174 | C50 |
| Prostate cancer | 185 | C61 |

Abbreviations: ICD, International Classification of Diseases; NMSC, non-melanoma skin cancer.

### Supplemental Table 2. Covariate adjusted differences in the 10^th^, 25^th^, 50^th^, 75^th^, and 90^th^ percentiles of the distributions of each neuroimaging outcome, comparing participants with versus without cancer history in quantile regression models.

|  |  |  |  | Demographics-Adjusted Base Model^b^ |  |  |  |  | Fully Adjusted Model^c^ |  |  |
| --- | --- | --- | --- | --- | --- | --- | --- | --- | --- | --- | --- |
| Outcome | N^a^ | Percentile of the outcome | Estimate | 95% CI | p-value | p-value for the Wald test |  | Estimate | 95% CI | p-value | p-value for the Wald test |
| GM | 41,378 | 10 | -697 | -2181 to 683 | 0.422 |  |  | -581 | -1660 to 761 | 0.490 |  |
|  |  | 25 | -621 | -1470 to 482 | 0.264 |  |  | -350 | -1237 to 791 | 0.614 |  |
|  |  | 50 | -647 | -1496 to 262 | 0.134 | 0.508 |  | -579 | -1331 to 312 | 0.192 | 0.715 |
|  |  | 75 | -557 | -1570 to 381 | 0.232 |  |  | -414 | -1280 to 562 | 0.420 |  |
|  |  | 90 | 353 | -884 to 1722 | 0.748 |  |  | 185 | -763 to 1644 | 0.590 |  |
| TBV | 41,378 | 10 | -1364 | -3041 to 589 | 0.220 |  |  | -1384 | -3274 to 614 | 0.196 |  |
|  |  | 25 | -938 | -2395 to 694 | 0.284 |  |  | -902 | -2624 to 957 | 0.334 |  |
|  |  | 50 | -986 | -2247 to 690 | 0.332 | 0.497 |  | -631 | -2011 to 708 | 0.440 | 0.439 |
|  |  | 75 | 40 | -1305 to 1656 | 0.820 |  |  | -333 | -1896 to 1265 | 0.832 |  |
|  |  | 90 | -321 | -2091 to 2154 | 0.894 |  |  | 149 | -1823 to 2191 | 0.960 |  |
| HV | 42,036 | 10 | -23 | -52 to 8 | 0.130 |  |  | -22 | -52 to 13 | 0.258 |  |
|  |  | 25 | -5 | -31 to 15 | 0.600 |  |  | -3 | -25 to 20 | 0.898 |  |
|  |  | 50 | -17 | -41 to 3 | 0.092 | 0.253 |  | -9 | -30 to 11 | 0.424 | 0.261 |
|  |  | 75 | -30 | -56 to -6 | 0.008 |  |  | -26 | -54 to -1 | 0.046 |  |
|  |  | 90 | -31 | -59 to 1 | 0.072 |  |  | -24 | -55 to 10 | 0.198 |  |
| WMH | 40,119 | 10 | -43 | -121 to 22 | 0.132 |  |  | -49 | -112 to 19 | 0.150 |  |
|  |  | 25 | -30 | -90 to 30 | 0.368 |  |  | -24 | -105 to 35 | 0.428 |  |
|  |  | 50 | 40 | -50 to 136 | 0.422 | 0.008 |  | 35 | -51 to 125 | 0.424 | <0.001 |
|  |  | 75 | 258 | 69 to 445 | 0.008 |  |  | 250 | 71 to 446 | 0.014 |  |
|  |  | 90 | 524 | 172 to 981 | <0.001 |  |  | 552 | 250 to 1002 | 0.002 |  |
| CTAD | 42,036 | 10 | -0.005 | -0.012 to -0.001 | 0.080 |  |  | -0.006 | -0.011 to -0.000 | 0.026 |  |
|  |  | 25 | -0.004 | -0.008 to -0.000 | 0.018 |  |  | -0.002 | -0.007 to 0.002 | 0.130 |  |
|  |  | 50 | -0.005 | -0.009 to -0.002 | <0.001 | 0.012 |  | -0.005 | -0.009 to -0.001 | 0.002 | 0.003 |
|  |  | 75 | -0.004 | -0.007 to 0.001 | 0.054 |  |  | -0.002 | -0.007 to 0.002 | 0.140 |  |
|  |  | 90 | 0.002 | -0.003 to 0.007 | 0.224 |  |  | 0.003 | -0.003 to 0.007 | 0.188 |  |

Abbreviations: CI, confidence interval; GM, gray matter; TBV, total brain volume; HV, hippocampal volume; WMH, white matter hyperintensity; AD, Alzheimer’s disease; CTAD, cortical thickness in the AD signature region.

^a^ Number of participants included in the analyses per outcome. Standard errors were obtained using a nonparametric bootstrap with the individual as the independent unit to account for a small number of repeated MRIs.

^b^ Adjusted for age, sex, race, and APOE-ε4 carrier status.

^c^ Further adjusted for education and Townsend deprivation index, BMI, ever smoking, ever using alcohol, physical activity, and assessment center.

### Supplementary Table 3. Association of site-specific cancer diagnosis with neuroimaging outcomes: β coefficients from linear mixed regression models.

|  |  |  | Demographics-Adjusted Base Model^b^ |  |  |  | Fully Adjusted Model^c^ |  |
| --- | --- | --- | --- | --- | --- | --- | --- | --- |
| Outcome | N^a^ | Estimate | 95% CI | p-value |  | Estimate | 95% CI | p-value |
| ***NMSC (n = 2,392)*** | | | | | | | | |
| GM | 37,029 | 931 | -72 to 1934 | 0.069 |  | 569 | -452 to 1590 | 0.275 |
| TBV | 37,029 | 1236 | -371 to 2843 | 0.132 |  | 847 | -800 to 2495 | 0.313 |
| HV | 37,624 | -26 | -50 to -1 | 0.038 |  | -24 | -49 to 2 | 0.066 |
| WMH | 35,893 | 57 | -197 to 311 | 0.661 |  | 109 | -149 to 366 | 0.409 |
| CTAD | 37,624 | 0.000 | -0.005 to 0.004 | 0.959 |  | 0.000 | -0.004 to 0.005 | 0.938 |
| ***Breast cancer^d^ (n = 1,087)*** | | | | | | | | |
| GM | 37,095 | 162 | -1327 to 1651 | 0.831 |  | -332 | -1853 to 1189 | 0.669 |
| TBV | 37,095 | -3112 | -5502 to -721 | 0.011 |  | -3450 | -5908 to -993 | 0.006 |
| HV | 37,686 | -32 | -69 to 5 | 0.093 |  | -32 | -70 to 6 | 0.098 |
| WMH | 35,954 | 567 | 194 to 939 | 0.003 |  | 512 | 132 to 891 | 0.008 |
| CTAD | 37,686 | -0.007 | -0.013 to 0.000 | 0.055 |  | -0.008 | -0.014 to -0.001 | 0.031 |
| ***Prostate cancer^e^ (n = 825)*** | | | | | | | | |
| GM | 36,877 | -929 | -2576 to 717 | 0.269 |  | -687 | -2369 to 995 | 0.423 |
| TBV | 36,877 | 2875 | 233 to 5518 | 0.033 |  | 2832 | 114 to 5551 | 0.041 |
| HV | 37,473 | 0 | -40 to 39 | 0.986 |  | -1 | -42 to 40 | 0.951 |
| WMH | 35,741 | 392 | -27 to 811 | 0.067 |  | 252 | -174 to 679 | 0.246 |
| CTAD | 37,473 | -0.004 | -0.012 to 0.003 | 0.244 |  | -0.004 | -0.011 to 0.004 | 0.342 |

Abbreviations: CI, confidence interval; NMSC, non-melanoma skin cancer; GM, gray matter; TBV, total brain volume; HV, hippocampal volume; WMH, white matter hyperintensity; AD, Alzheimer’s disease; CTAD, cortical thickness in the AD signature region.

^a^ Number of participants included in the analyses per outcome. Mixed effects models with random intercepts for individuals were used to account for a small number of repeated MRIs.

^b^ Adjusted for age, sex, race, and APOE-ε4 carrier status.

^c^ Further adjusted for education and Townsend deprivation index, BMI, ever smoking, ever using alcohol, physical activity, and assessment center.

^d^ Among females only.

^e^ Among males only.

### Supplementary Table 4. Association of cancer diagnosis with neuroimaging outcomes with a 5-year washout period: β coefficients from linear mixed regression models.

|  |  |  | Demographics-Adjusted Base Model^b^ |  |  |  | Fully Adjusted Model^c^ |  |
| --- | --- | --- | --- | --- | --- | --- | --- | --- |
| Outcome | N^a^ | Estimate | 95% CI | p-value |  | Estimate | 95% CI | p-value |
| GM | 40,086 | -101 | -904 to 701 | 0.804 |  | 46 | -769 to 862 | 0.911 |
| TBV | 40,086 | 8 | -1275 to 1292 | 0.990 |  | 43 | -1269 to 1355 | 0.949 |
| HV | 40,723 | -24 | -44 to -3 | 0.021 |  | -19 | -40 to 2 | 0.073 |
| WMH | 38,870 | 174 | -27 to 374 | 0.090 |  | 153 | -51 to 357 | 0.142 |
| CTAD | 40,723 | -0.002 | -0.005 to 0.002 | 0.347 |  | -0.001 | -0.005 to 0.002 | 0.540 |

Abbreviations: CI, confidence interval; GM, gray matter; TBV, total brain volume; HV, hippocampal volume; WMH, white matter hyperintensity; AD, Alzheimer’s disease; CTAD, cortical thickness in the AD signature region.

^a^ Number of participants included in the analyses per outcome. Mixed effects models with random intercepts for individuals were used to account for a small number of repeated MRIs.

^b^ Adjusted for age, sex, race, and APOE-ε4 carrier status.

^c^ Further adjusted for education and Townsend deprivation index, BMI, ever smoking, ever using alcohol, physical activity, and assessment center.

### Supplementary Table 5. Association of cancer diagnosis with neuroimaging outcomes based on time between cancer diagnosis and MRI scan: β coefficients from linear mixed regression models.

|  |  |  |  | Demographics-Adjusted Base Model^b^ |  |  |  | Fully Adjusted Model^c^ |  |
| --- | --- | --- | --- | --- | --- | --- | --- | --- | --- |
| Outcome | N^a^ | Group | Estimate | 95% CI | p-value |  | Estimate | 95% CI | p-value |
| GM | 41,378 | No cancer | Ref. |  | Ref. |  | Ref. |  | Ref. |
|  |  | 0-1 years | -452 | -2983 to 2079 | 0.726 |  | -693 | -3292 to 1906 | 0.601 |
|  |  | 1-5 years | -1538 | -2792 to -284 | 0.016 |  | -1516 | -2797 to -236 | 0.020 |
|  |  | 5-10 years | -92 | -1490 to 1306 | 0.898 |  | 16 | -1411 to 1443 | 0.983 |
|  |  | >10 years | -235 | -1356 to 886 | 0.681 |  | -196 | -1341 to 948 | 0.737 |
| TBV | 41,378 | No cancer | Ref. |  | Ref. |  | Ref. |  | Ref. |
|  |  | 0-1 years | -922 | -4951 to 3106 | 0.654 |  | -1903 | -6079 to 2274 | 0.372 |
|  |  | 1-5 years | -594 | -2593 to 1404 | 0.560 |  | -831 | -2888 to 1227 | 0.429 |
|  |  | 5-10 years | -476 | -2704 to 1753 | 0.676 |  | -536 | -2830 to 1758 | 0.647 |
|  |  | >10 years | -594 | -2385 to 1197 | 0.516 |  | -599 | -2440 to 1241 | 0.523 |
| HV | 42,036 | No cancer | Ref. |  | Ref. |  | Ref. |  | Ref. |
|  |  | 0-1 years | -7 | -65 to 50 | 0.803 |  | -3 | -62 to 57 | 0.932 |
|  |  | 1-5 years | -21 | -50 to 9 | 0.166 |  | -16 | -46 to 14 | 0.295 |
|  |  | 5-10 years | -38 | -71 to -6 | 0.022 |  | -35 | -68 to -1 | 0.044 |
|  |  | >10 years | -29 | -57 to -1 | 0.041 |  | -28 | -57 to 0 | 0.051 |
| WMH | 40,119 | No cancer | Ref. |  | Ref. |  | Ref. |  | Ref. |
|  |  | 0-1 years | -46 | -741 to 650 | 0.898 |  | -119 | -817 to 579 | 0.738 |
|  |  | 1-5 years | 466 | 130 to 803 | 0.007 |  | 391 | 51 to 731 | 0.024 |
|  |  | 5-10 years | 507 | 135 to 879 | 0.008 |  | 373 | -2 to 749 | 0.052 |
|  |  | >10 years | 45 | -240 to 330 | 0.757 |  | 84 | -206 to 374 | 0.568 |
| CTAD | 42,036 | No cancer | Ref. |  | Ref. |  | Ref. |  | Ref. |
|  |  | 0-1 years | 0.000 | -0.013 to 0.012 | 0.970 |  | 0.002 | -0.011 to 0.014 | 0.799 |
|  |  | 1-5 years | -0.013 | -0.019 to -0.007 | <0.001 |  | -0.012 | -0.018 to -0.006 | 0.001 |
|  |  | 5-10 years | -0.006 | -0.012 to 0.001 | 0.088 |  | -0.004 | -0.011 to 0.003 | 0.243 |
|  |  | >10 years | -0.002 | -0.007 to 0.003 | 0.446 |  | -0.002 | -0.007 to 0.003 | 0.443 |

Abbreviations: CI, confidence interval; GM, gray matter; TBV, total brain volume; HV, hippocampal volume; WMH, white matter hyperintensity; AD, Alzheimer’s disease; CTAD, cortical thickness in the AD signature region.

^a^ Number of participants included in the analyses per outcome. Mixed effects models with random intercepts for individuals were used to account for a small number of repeated MRIs.

^b^ Adjusted for age, sex, race, and APOE-ε4 carrier status.

^c^ Further adjusted for education and Townsend deprivation index, BMI, ever smoking, ever using alcohol, physical activity, and assessment center.

### Supplementary Table 6. Association of cancer diagnosis with neuroimaging outcomes adjusting for selection using IPW: β coefficients from weighted linear mixed regression models.

|  |  |  | Demographics-Adjusted Base Model^b^ |  |  |  | Fully Adjusted Model^c^ |  |
| --- | --- | --- | --- | --- | --- | --- | --- | --- |
| Outcome | N^a^ | Estimate | 95% CI | p-value |  | Estimate | 95% CI | p-value |
| GM | 40,943 | -472 | -1172 to 229 | 0.187 |  | -364 | -1075 to 348 | 0.314 |
| TBV | 40,943 | -282 | -1402 to 839 | 0.622 |  | -380 | -1525 to 765 | 0.515 |
| HV | 41,593 | -23 | -40 to -5 | 0.010 |  | -19 | -36 to -1 | 0.039 |
| WMH | 39,702 | 222 | 39 to 405 | 0.017 |  | 185 | 0 to 370 | 0.051 |
| CTAD | 41,593 | -0.005 | -0.008 to -0.002 | 0.003 |  | -0.004 | -0.007 to -0.001 | 0.011 |

Abbreviations: IPW, inverse probability weighting; CI, confidence interval; GM, gray matter; TBV, total brain volume; HV, hippocampal volume; WMH, white matter hyperintensity; AD, Alzheimer’s disease; CTAD, cortical thickness in the AD signature region.

^a^ Number of participants included in the analyses per outcome. Mixed effects models with random intercepts for individuals were used to account for a small number of repeated MRIs.

^b^ Adjusted for age, sex, race, and APOE-ε4 carrier status.

^c^ Further adjusted for education and Townsend deprivation index, BMI, ever smoking, ever using alcohol, physical activity, and assessment center.
